# Diet quality and obesity in women of reproductive age in Northern Tanzania: a cross-sectional study

**DOI:** 10.1101/2025.07.29.25332361

**Authors:** Lilia Bliznashka, Fusta Azupogo, Elise Reynolds, Charles D Arnold, Sonja Y Hess, Joyce Kinabo, Kidola Jeremiah, Evangelista Malindisa, Deanna K Olney, Marie T Ruel

## Abstract

**Background:** Obesity is an increasing problem among women of reproductive age (WRA) in Tanzania.

**Objective:** We described WRA’s nutritional status by socio-demographic factors and assessed associations with diet quality.

**Methods:** We analysed baseline data from a cluster-randomised controlled trial in Arusha and Kilimanjaro regions (n=2,415). Diet was assessed using a quantitative 24-hour recall. We calculated the Global Diet Quality Score (GDQS; 0-49), with higher scores indicating healthier diet. General obesity was defined as body mass index (BMI)≥30 kg/m^2^; morbid obesity as BMI≥35 kg/m^2;^; and central obesity as: waist circumference (WC)≥80 cm, WC≥88 cm, waist-to-hip ratio (WHR)≥0.85, waist-to-height ratio (WHtR)≥0.50, and WHR≥0.85 or BMI≥30 kg/m^2^. We tested associations between diet quality and nutritional status using generalised linear models controlling for age and sociodemographic factors and tested interactions to assess differential associations by age groups.

**Results:** The prevalence of general obesity was 25.1%, morbid obesity 8.4%, and central obesity 48.2-71.6% depending on the definition. Mean GDQS was 20.9±3.9. General and central obesity were more prevalent among women who were older, less educated, had light physical labour occupations, were in the highest wealth quintile, and lived in more urbanised villages and in more food secure households. Higher GDQS was associated with lower risk of morbid obesity: risk ratio (RR) 0.97 (95% CI 0.94, 1.00). Higher GDQS was also associated with 0.25-0.27 kg/m^2^ lower BMI, 0.54-0.66 cm lower WC, and 0.53-0.58 cm lower hip circumference in women 30-49 years of age.

**Conclusion:** Better diet quality emerged as a protective factor for morbid obesity and for other obesity measures among women 30-49 years of age. Our study suggests that interventions to improve diet quality in Tanzania should target women in their thirties and forties and those with lower physical activity and higher education, food security, and wealth to maximise effectiveness. **Keywords:** obesity, overweight, BMI, Tanzania, women of reproductive age, low- and middle-income countries

## Introduction

Obesity is a major public health concern globally. Obesity prevalence in women has been increasing globally, with particularly high increases in low- and middle-income countries (LMICs), including in some countries in sub-Saharan Africa.^1,2^ In Tanzania, overweight and obesity prevalence in adult women increased from 26.1% in 1990 to 44.7% in 2021.^2^ In 2021, five million Tanzanian women were overweight and obese and this number is projected to increase to 17.7 million by 2050 (or 62.6% of adult women).^2^ Among Tanzanian women of reproductive age (WRA, 15-49 years of age), overweight and obesity increased by 30% in less than a decade from 28% in 2015-16 to 36% in 2021-22.^3,4^

Body mass index (BMI) is widely used to measure general obesity, defined as BMI≥30 kg/m^2^.^5^ Obesity increases the risk of cardiovascular morbidity and mortality.^6^ In WRA, obesity is also associated with poor perinatal health, including gestational diabetes, pregnancy-induced hypertension, pre-eclampsia, and postpartum morbidity.^7,8^ BMI, however, does not discriminate between fat and lean mass, nor does it account for body fat distribution.^9,10^ Measures of central obesity – waist circumference (WC), waist-to-hip ratio (WHR), and waist-to-height ratio (WHtR), which capture the accumulation of fat around the abdominal area – are better predictors of cardiometabolic morbidity and mortality than general obesity.^11–13^ Using multiple metrics to describe obesity is likely to provide a more nuanced understanding of the obesity burden in Tanzanian WRA.

Unhealthy diets are a well-established risk factor for overweight and obesity, together with reduced physical activity and sedentary lifestyle, alcohol use, and smoking.^14^ Although the relationship between unhealthy diets and overweight and obesity is well-established in high-income countries,^14–16^ less is known in LMICs where research has focused on data-derived dietary patterns, rather than dietary intake data, and overweight and obesity.^17–20^ The Global Diet Quality Score (GDQS) was designed to measure consumption of healthy food groups and avoidance of unhealthy food groups and to predict both undernutrition and diet-related non-communicable diseases (NCDs) risks.^21^ Since its development, studies have demonstrated that higher GDQS (i.e., higher diet quality) is associated with less underweight among WRA in rural sub-Saharan Africa.^22,23^ However, research on the association between GDQS and overweight and obesity is more limited and inconsistent.^24^ In Tanzania, one study showed that higher diet quality, measured by the Prime Dietary Quality Score (PDQS, a precursor to GDQS) was associated with lower odds of overweight among women living in Dar es Salaam.^25^ Evidence on whether diet quality is associated with women’s nutritional status in rural contexts or other regions in Tanzania is lacking. Further examining and comparing associations with measures of general and central obesity can improve our understanding of potential dietary changes that could help control the expected rise in obesity in Tanzania over the next few decades.

The objectives of this paper were (1) to describe the nutritional status of WRA in Tanzania, with a focus on overweight and obesity, and (2) to assess the associations between diet quality (measured by GDQS) and overweight, general obesity, and central obesity. We hypothesised that higher diet quality would be associated with a lower risk of overweight and obesity, with no *a priori* hypotheses on which overweight and obesity metrics would be more strongly associated with diet quality. We also described women’s nutritional status by sociodemographic characteristics (age, education, wealth, food security, urbanicity, and employment) to identify potential indicators for targeting obesity prevention.

## Methods

### Participants

We used baseline data from an on-going cluster randomised controlled trial being implemented in 33 villages in five districts in Arusha and Kilimanjaro regions in Northern Tanzania. The trial was designed to assess the effect of a package of supply, demand, and food environment interventions on household vegetable production and fruit and vegetable intake among WRA.^26^ Women were eligible for inclusion if they were 15-49 years of age, had at least one biological child 10-14 years of age living with them, and planned to remain in the study area for the duration of the trial. Prior to enrolment, listings of all eligible households in each village were obtained from village and hamlet leaders. Households were randomly selected from these household listings for trial inclusion. The trial enrolled 2,604 WRA between October 11, 2023 and January 15, 2024. An average of 102 WRA were enrolled per village, ranging from 15 to 211. Data were collected by trained enumerators using quantitative questionnaires. All enumerators received a three-week training prior to the start of data collection.

### Data collection

Trained enumerators measured height, weight, hip and waist circumference of WRA who wore lightweight clothing and no shoes for the assessment. Height was measured to the nearest 0.1 cm using a mechanical stadiometer (SECA213); weight was measured to the nearest 0.1 kg using a digital scale (SECA876); hip and waist circumference were measured using a mechanical tape (SECA203). All measurements were taken in duplicate. The average of the two was used in the analysis.

BMI was calculated as weight (kg) divided by height (m) squared. Women were categorised as having underweight (BMI<18.5 kg/m^2^), normal weight (BMI≥18.5 kg/m^2^ and BMI<25 kg/m^2^), overweight (BMI≥25 kg/m^2^ and BMI<30 kg/m^2^), obesity (BMI≥30 kg/m^2^ and

BMI<35 kg/m^2^), or morbid obesity (BMI ≥35 kg/m^2^).^5,27^ Any overweight/obesity was defined as BMI≥25 kg/m^2^. General obesity was defined as BMI≥30 kg/m^2^. We used five definitions of central obesity: WC≥80 cm, WC≥88 cm, WHR≥0.85, WHtR≥0.50, and WHR≥0.85 or BMI≥30 kg/m^2^.^28–30^ The continuous outcomes of BMI, WC, and hip circumference were also used in the analysis.

Women’s diet was assessed using a quantitative multi-pass 24-hour recall.^31^ Standard recipes were used for estimation of mixed dishes. Details on data collection, intake estimation, and recipe formulation for mixed dishes are available elsewhere.^32^ Data collection was paused from December 20, 2023 to January 7, 2024 to minimise recall bias associated with the Christmas and New Year holiday food consumption. GDQS was derived from the 24-hour recall (single day) following the recommended guidelines.^21,33^ We calculated total GDQS (range 0 to 49) based on 25 food groups, and its 2 subcomponents: GDQS+ (range 0 to 32) based on 16 healthy food groups and GDQS- (range 0 to 17) based on 7 unhealthy food groups and 2 food groups considered unhealthy if consumed above a specified threshold.^33^ Higher scores are given to higher consumption of healthy food groups (GDQS+) and to lower consumption of unhealthy food groups (GDQS-).

Household food security was assessed using the Food Insecurity Experience Scale.^34^ We calculated total score (range 0-8), and categorised households as food secure (score=0), mildly food insecure (1≤score≤3), moderately food insecure (4≤score≤6), or severely food insecure (score≥7).^35^ A household wealth index was calculated using principal components analysis of 12 assets and seven housing characteristics and divided into quintiles. We developed a village urbanicity scale^36^ and divided the scale into tertiles, which we interpreted as rural villages (lowest tertile), less urbanised villages (middle tertile), and more urbanised villages (highest tertile). Women’s occupation was used as a proxy for physical activity. Occupation was divided into moderate/heavy physical labour required (i.e., self-employed farming, self-employed animal husbandry, agriculture wage labour, animal husbandry wage labour) or light physical labour required (i.e., enterprise/business, salaried government, salaried private sector, non-agricultural wage labour, household work, food retail, food collection, or other). Women who reported being unemployed or not currently seeking work were classified as having light physical labour occupation.

### Statistical analysis

Of the 2,604 WRA enrolled, 2,598 (99.8%) women had diet data. Consistent with other analyses of the trial population, we excluded women with total daily energy intake <1^st^ percentile (< 500 kcal/day) or >99^th^ percentile (> 6,000 kcal/day) of the distribution (n=49, 1.9%).^32^ We also excluded pregnant WRA (n=121, 4.6%), since their diets may be different, and we did not collect data on trimester of pregnancy or gestational weight gain and were thus unable to adjust nutritional status metrics for pregnancy status. Thirteen additional WRA (0.5%) were excluded due to missing BMI, WC, or hip circumference data. The final analytic sample contained 2,415 WRA. We compared WRA included in the analytic sample to those excluded using t-tests, considering differences significant at p<0.05.

Of these 2,415 WRA in the analytic sample, 20% (n=482) were lactating WRA. We did not collect data on number of months postpartum or age of the breastfed child. As a result, we could not adjust nutritional status metrics for lactation status or postpartum weight loss. Therefore, as a sensitivity analyses, we reran all analyses excluding lactating WRA from the analytic sample.

We used descriptive statistics (mean and SD for continuous variables and proportion for binary variables) to describe WRA’s nutritional status (objective 1). We examined statistics for the full sample and disaggregated them by region, urbanicity (rural vs less urbanised vs more urbanised villages), woman’s age (20-29, 30-39, 40-49 years), woman’s education (none complete vs primary/higher), woman’s occupation (moderate/heavy vs light physical labour required), household wealth quintile, and household food security category. Missing data on covariates (0.3% for household wealth) were imputed using mean village-level imputation.

We assessed the associations between diet quality and overweight and obesity using bivariate and multivariable generalised linear models (objective 2). Multivariable models controlled for woman’s age, education and occupation, household wealth, region, and urbanicity. We did not control for daily energy intake or household food insecurity because they could be on the causal pathway between GDQS and nutritional status.^37^ We used log-binomial models for binary outcomes to estimate adjusted risk ratios (RR) and 95% CIs. When a log-binomial model did not converge, we used a log-Poisson model which estimates unbiased RR for common outcomes but with less precision than log-binomial models.^38^ We used linear models for continuous outcomes to estimate adjusted mean differences (MD) and 95% CIs. Given that overweight and obesity increase with age,^4^ we further explored whether the associations between diet quality and overweight and obesity differed by woman’s age group (20-29, 30-39, 40-49 years). We included an interaction term between total GDQS and woman’s age group, and estimated associations within age subgroups. Interactions were considered significant at p<0.05. In all models, standard errors were clustered at the village level in line with the trial design. Models did not adjust for intervention assignment. Given the exploratory nature of the analysis, we did not adjust for multiple hypothesis testing. All analyses were conducted in Stata 18.

### Ethical considerations

Ethical approval was received by the National Health Research Ethics Committee of the National Institute of Medical Research in Tanzania (NIMR/HQ/R.8a/Vol.IX/4357), the Institutional Review Board of International Food Policy Research Institute (#00007490), and the Research Ethics Committee of Wageningen University and Research (#2023-022). Trained enumerators provided information about the study. Written informed consent was obtained from the household head and the WRA.

## Results

### Sample descriptives

Women were 38.4±6.2 years old on average, and 81.4% had completed primary or higher education (**Table 1**). Nearly two-thirds of women were involved in occupations requiring moderate/heavy physical labour. One-fifth of women were lactating, and 15.6% lived in female-headed households. On average, women consumed 2,400 kcal/day. Total GDQS was 20.9±3 on average (out of 49 points), with GDQS+ (healthy food groups) contributing 8.8±3.3 points (out of 32 points) and GDQS-(unhealthy food groups) contributing 12.1±1.9 (out of 17 points) on average (Table 1). Scores for individual food groups comprising GDQS, the number of food items by GDQS group, the proportion of women who scored zero on each food group, and the proportion who scored the highest on each food group are shown in **Supplemental Table 1.** More than 94% of women scored zero on citrus fruit, deep orange fruit, other fruit, deep orange tubers, nuts and seeds, poultry and game meet, low-fat dairy, eggs, and red meat (reflecting low or no consumption). Over 20% of women reported consuming refined grains and baked goods, sweets and ice cream, sugar-sweetened beverages, and deep-fried foods in harmful amounts (Supplemental Table 1).

**Table 1.** Household and individual characteristics of the 2,415 women of reproductive age (15-49 years of age) included in the analytic sample.

| | Mean ± SD or % |
| --- | --- |
| N | 2,415 |
| *Household characteristics* |  |
| Size | 5.8±1.7 |
| Household head age | 46.8±9.9 |
| Household head is female | 15.6 |
| Household head has completed primary/higher education | 82.5 |
| *Woman characteristics* |  |
| Age (in years) | 38.4±6.2 |
| Age group |  |
| 20-29 years | 7.6 |
| 30-39 years | 47.0 |
| 40-49 years | 45.4 |
| Has completed primary/higher education | 81.4 |
| Occupation |  |
| Light physical labour required | 36.1 |
| Moderate/heavy physical labour required | 63.9 |
| Lactating | 20.0 |
| *Women's diet* |  |
| Current energy intake (kcal/day) | 2,400.3±1,018.0 |
| Global Diet Quality Score (GDQS) (range 0-49) | 20.9±3.9 |
| GDQS+ (range 0-32) | 8.8±3.3 |
| GDQS- (range 0-17) | 12.1±1.9 |

Overall, women included in the analytic sample were significantly older (by ∼2 years) than women excluded from the analytic sample due to missing data on diet or anthropometry, pregnancy, or very high/low daily energy intake (**Supplemental Table 2**). Similarly, household heads were ∼2 years older on average in the sample included in the analysis compared to the excluded sample. The two samples were similar on all other characteristics including woman’s or head of household education, woman’s occupation, and household size.

### WRA nutritional status

Average BMI in our WRA sample was 26.5±6.5 kg/m^2^, with 4.8% of women underweight, and 41.6% of normal weight (**Table 2**). More than half of the sampled women (53.6%) were overweight or obese (BMI ≥25 kg/m^2^) and one quarter (25.1%) suffered from general obesity (BMI≥30 kg/m^2^). The prevalence of central obesity was high and varied depending on the definition, ranging from 48.2% (WC≥88 cm) to a very high of 71.6% (WHtR≥0.5). Nutritional status was similar among the 1,933 non-pregnant and non-lactating women (**Supplemental Table 3**).

**Table 2.** Nutritional status of the 2,415 women of reproductive age (15-49 years of age) included in the analytic sample.

| | Mean ± SD or<br>% |
| --- | --- |
| N | 2,415 |
| Weight (kg) | 66.9±15.8 |
| Height (cm) | 158.9±6.5 |
| Body mass index (BMI, kg/m<sup>2</sup>) | 26.5±6.0 |
| Hip circumference | 102.9±12.3 |
| Waist circumference | 88.1±13.3 |
| *BMI categories* |  |
| Underweight (BMI <18.5 kg/m<sup>2</sup>) | 4.8 |
| Normal weight (BMI 18.5-24.9 kg/m<sup>2</sup>) | 41.6 |
| Overweight (BMI 25-29.9 kg/m<sup>2</sup>) | 28.5 |
| Obesity (BMI 30-34.9 kg/m<sup>2</sup>) | 16.6 |
| Morbid obesity (BMI≥35 kg/m<sup>2</sup>) | 8.4 |
| Any overweight/obesity (BMI≥25 kg/m<sup>2</sup>) | 53.6 |
| General obesity (BMI≥30 kg/m<sup>2</sup>) | 25.1 |
| *Central obesity* |  |
| Waist circumference ≥80 cm | 71.1 |
| Waist circumference ≥88 cm | 48.2 |
| Waist-to-hip ratio ≥0.85 | 53.6 |
| Waist-to-height ratio ≥0.5 | 71.6 |
| Waist-to-hip ratio≥0.85 or BMI≥30 kg/m<sup>2</sup> | 60.4 |

Obesity prevalence (measured by several anthropometric measures) increased sharply from age 20-29 to 30-39 years, with little change thereafter (**Figure 1, Supplemental Table 4**). Obesity was more prevalent in women with primary or higher education than women without completed education. With respect to occupation, obesity was more prevalent among women with light physical labour occupations than women with moderate/heavy physical labour occupations. Conversely, underweight and normal weight were more prevalent in younger women 20-29 years of age, less educated women, and those whose occupation required moderate/heavy physical labour (Supplemental Table 4).

**Figure 1.**
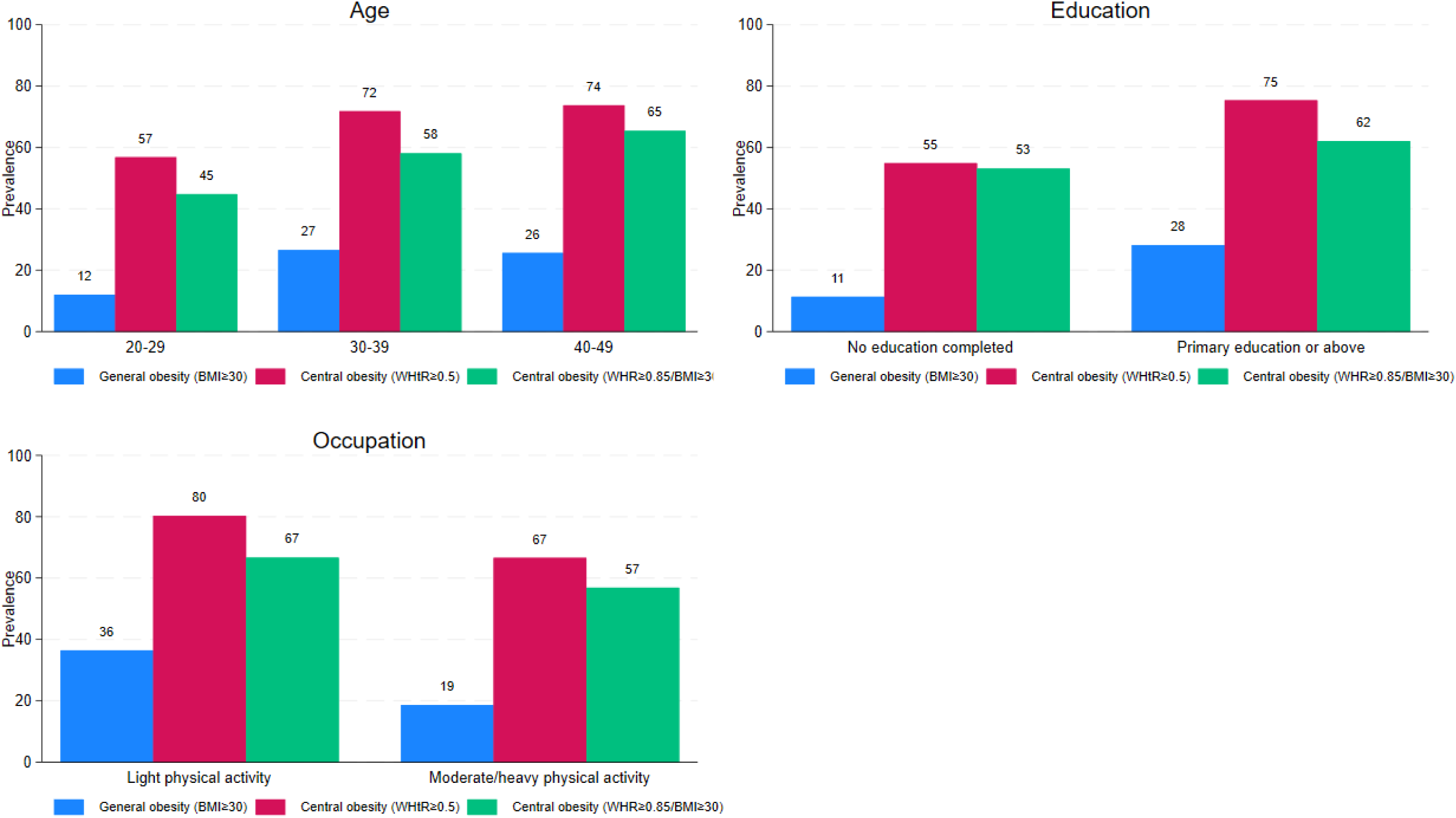
Obesity prevalence by metric and by woman’s age, education, and occupation in 2,415 women of reproductive age (15-49 years of age) included in the analytic sample **Abbreviations used:** BMI, body mass index; WHtR, waist-to-height ratio; WHR, waist-to-hip ratio.

When examining nutritional status by household wealth quintile, we observed that all measures of overweight and obesity increased with household wealth and underweight and normal weight decreased (**Figure 2**, Supplemental Table 4). Between 44.3% and 87.9% of women in the highest wealth quintile were obese, depending on the definition. With respect to food insecurity, overweight and obesity were more prevalent among food secure households and less prevalent in severely food insecure households, whereas the reverse was observed for underweight (**Figure 2**, Supplemental Table 4). However, this trend was less consistent among mildly and moderately food insecure households, where nutritional status was generally similar. Lastly, when considering urbanicity, we documented a gradient with higher prevalence of overweight and obesity with increasing urbanicity: from rural to less urbanised to more urbanised villages, regardless of the definition. The same gradient was observed for underweight and normal weight but in the opposite direction, with both more prevalent in rural villages than less urbanised or more urbanised ones.

**Figure 2.**
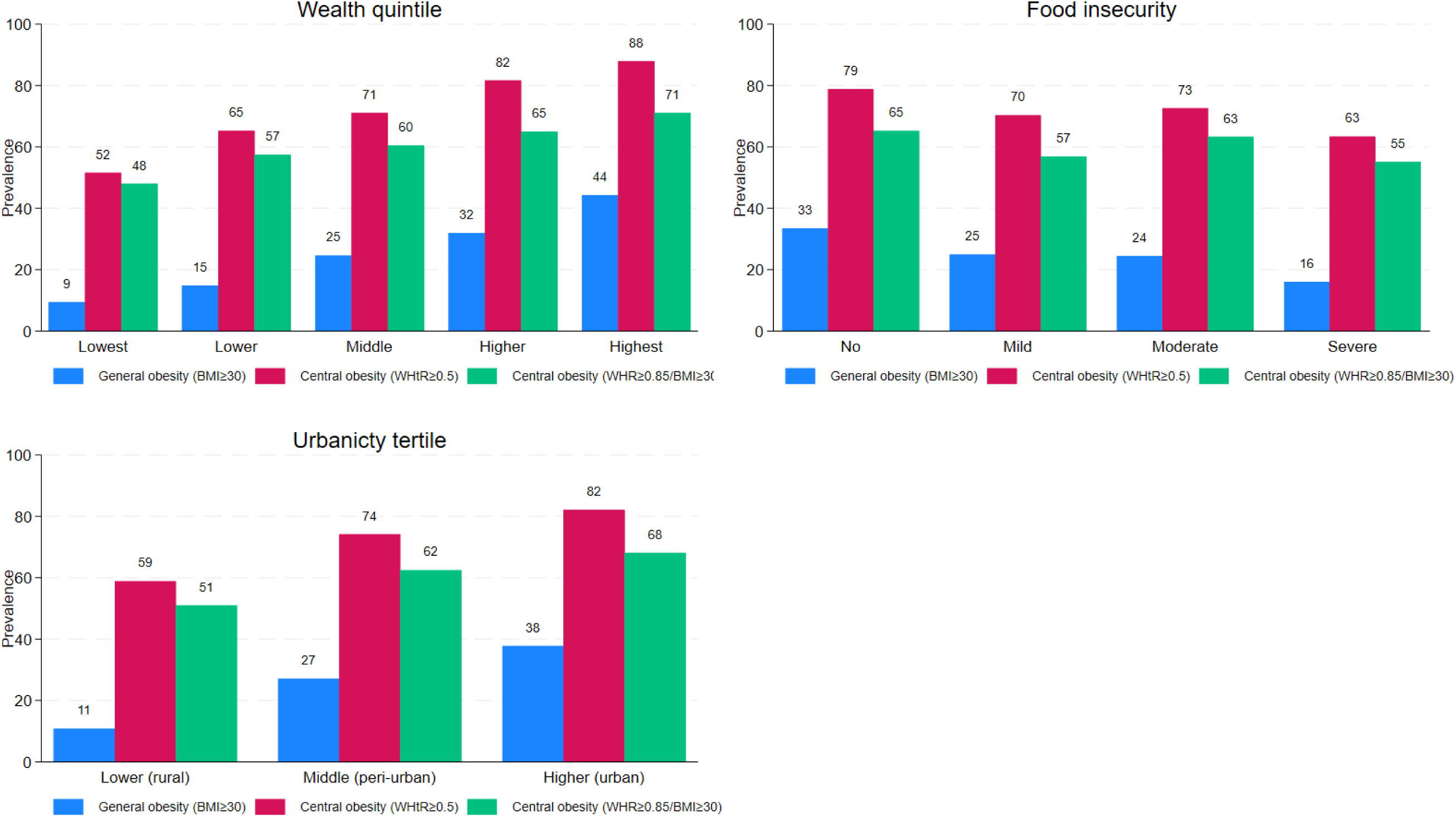
Obesity prevalence by metric and by household wealth, household food insecurity, and village urbanicity in 2,415 women of reproductive age (15-49 years of age) included in the analytic sample **Abbreviations used:** BMI, body mass index; WHtR, waist-to-height ratio; WHR, waist-to-hip ratio.

Overall, across the socio-economic characteristics we examined, the highest general obesity and central obesity prevalence were observed in women in the highest wealth quintile: 44.3% had BMI≥30 kg/m^2^ and 88.8% had WC≥80 cm and the highest prevalence of underweight was observed in the lowest wealth quintile (11.6%).

### Associations between diet quality and nutritional status

Total GDQS and GDQS+ were associated with a lower risk of morbid obesity: RR 0.97 (95% CI 0.94, 1.00) and RR 0.95 (95% CI 0.91, 0.99), respectively (**Figure 3**, **Supplemental Table 5**). GDQS and its sub-scores were not associated with other measures of obesity, underweight, or normal weight (Supplemental Tables 5). After excluding lactating women, GDQS and GDQS+ were no longer associated with morbid obesity (**Supplemental Table 6**). None of the GDQS scores were associated with BMI, waist or hip circumference in the full sample or in sensitivity analyses excluding lactating women (**Supplemental Tables 7 and 8**).

**Figure 3.**
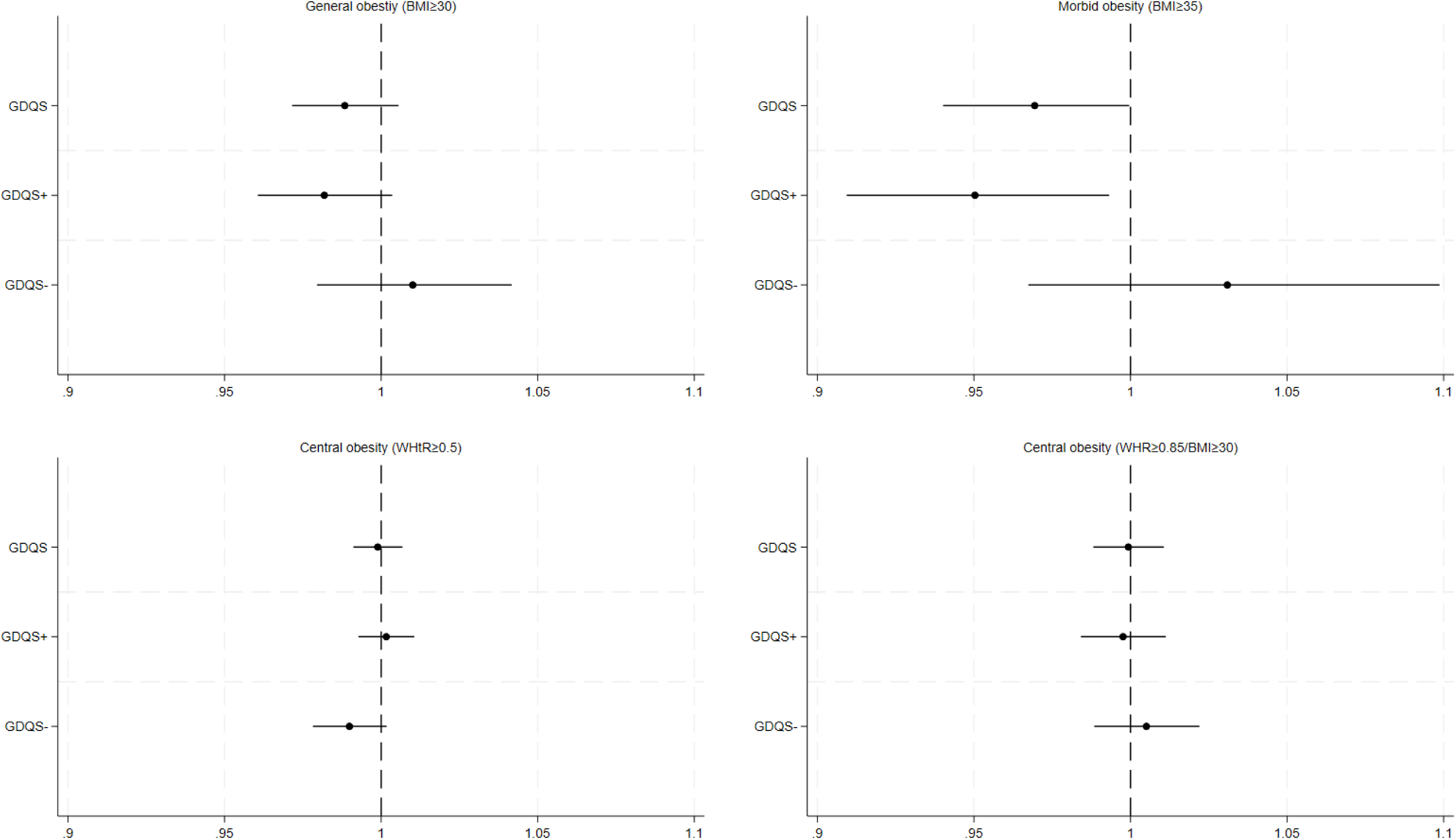
Multivariable adjusted associations between diet quality (GDQS, GDQS+ GDQS-) and obesity in 2,415 women of reproductive age (15-49 years of age) included in the analytic sample **Note:** Estimates are risk ratios (RR) and their 95% CIs from generalised linear models controlling for woman’s age, education, occupation, household wealth, region, and urbanicity. Standard errors were clustered at the village level. Abbreviations used: BMI, body mass index; GDQS, Global Diet Quality Score; WHtR, waist-to-height ratio; WHR, waist-to-hip ratio.

When examining whether the associations between diet quality and overweight and obesity varied by woman’s age, we found that total GQDS was associated with 8% lower risk of general obesity and 6% lower risk of WC≥88 cm in women 30-39 years of age, but not in women 20-29 or 40-49 years of age (**Supplemental Table 9**). In addition, total GDQS was associated with 0.25-0.27 kg/m^2^ lower BMI, 0.54-0.66 cm lower WC, and 0.53-0.58 lower hip circumference in women 30-39 years old and 40-49 years old, and with 0.19 kg/m^2^ higher BMI and 0.48 cm higher WC in women 20-29 years old.

## Discussion

In this study, we examined the nutritional status of WRA in Northern Tanzania and the associations between diet quality (measured by GDQS) and overweight and obesity. We documented a high prevalence of overweight and obesity, particularly among WRA in wealthier households and more urbanised villages. Underweight prevalence was generally low (<5%), with the lowest prevalence in the poorest households. The mean GDQS indicated a moderate risk of poor diet quality, characterised by inadequate intake of healthy foods and relatively low intake of unhealthy foods. We found no association between GDQS and underweight and limited evidence of an association with morbid obesity, but no association with any other measures of overweight or obesity. However, associations differed by woman’s age with higher GDQS having protective associations with general obesity, BMI, waist and hip circumference in women 30-39 and 40-49 years old. In younger women 20-29 years old, where overweight and obesity prevalence were lower, BMI and WC increased with higher GDQS.

Our findings of a high prevalence of any overweight/obesity and general obesity are consistent with existing literature on WRA in Tanzania.^25,39–41^ We built on this literature by examining the prevalence of central obesity using several measures, all of which have been shown to be better predictors of cardiometabolic risk and morbidity than general obesity.^11–13^ A study among pastoralists in the Arusha region showed a higher prevalence of central obesity defined by WHR≥0.85 and WC≥88 cm than general obesity.^40^ Consistent with these findings, we found that central obesity prevalence was higher than general obesity prevalence across all subgroups we examined, indicating that relying on BMI alone to understand obesity burden in this population may substantially underestimate the problem. Our results showed high variability in quantifying the obesity problem, depending on the definition and metric, ranging from 25.1% of women with general obesity (BMI≥30 kg/m^2^) to 71.6% with central obesity (WHtR≥0.5). Notably, a few studies have shown that African women have less body fat at the same WC as white women, indicating that higher WC cut-offs may be more suitable for African populations.^29^ In contrast, evidence suggests that WHtR is a more reliable predictor of cardiometabolic risks and disease than WC or WHR because it is less affected by sex or ethnicity.^12,30,42^

Overall, we found the highest prevalence of general and central obesity among older women, more educated women, women with light physical activity occupations, women in the wealthiest households, those living in food secure households, and those living in more urbanised villages, results consistent with the nutrition transition.^43^ Our previous unpublished analysis revealed a progressive increase in energy (2,250 to 2,526 kcal/day) and fat (57.7 to 88.8 g/day) intake across household wealth quintiles, from the lowest to the highest, consistent with our finding of increasing prevalence of central obesity with wealth. In fact, added fat and oil intake exceeded national dietary recommendations by approximately 50% (41 g/day vs. the recommended 28 g/day).^44^ The higher obesity prevalence in more urbanised villages is consistent with trends in Tanzania, which show that people in urban areas consume more high-sugar prepared foods than traditional staples and fresh foods.^45^ In addition, the intake of energy-dense, processed and ultra-processed foods eaten away from home, which are usually more available in more urbanised settings, has also increased substantially in Tanzania over the past two decades.^46^ In our sample, consumption of ultra-processed foods was relatively limited with ultra-processed foods accounting for 5% of total energy intake and two-thirds of women reporting no intake of ultra-processed foods (unpublished results). Overall, the high energy intake, largely from staples and fats, combined with potentially low physical activity levels and high consumption of ultra-processed foods by some women may explain the substantial burden of overweight and obesity in this population.

Our findings build on the limited literature on the associations between diet quality and overweight and obesity in Tanzania. A study of non-pregnant women (mean age 30) living in Dar Es Salaam showed that being in the highest diet quality quintile (measured by PDQS), was associated with a 24% lower risk of overweight and obesity than being in the lowest diet quality quintile.^25^ Our study also found that higher diet quality was associated with a 3 to 5% lower risk of morbid obesity in the full sample, and with a 6 to 8% lower risk of general obesity in women 30-39 years old. We extend the literature by demonstrating that higher diet quality was also associated with 0.25-0.27 kg/m^2^ lower BMI, 0.54-0.66 cm lower WC, and 0.53-0.58 lower hip circumference among women 30-49 years old. Further, our study is among the first to generate evidence on the relationship between diet quality and overweight and obesity in a rural sample in Tanzania, unlike the previous study conducted in the largest urban centre of the country.^25^

We further expand the evidence base by examining associations between diet quality and measures of central obesity. Published literature on GDQS to date has focused on its associations with underweight or BMI categories,^22–24^ while evidence on associations with central obesity is lacking. In the study among pastoralists mentioned above, higher dietary diversity tertile was associated with lower general and central obesity in women, albeit the latter was only significant when using the WHR≥0.85 definition and not the WC≥88 cm definition.^40^ More importantly, this study used a household dietary diversity score, which is a proxy for food access^47^ rather than quality of individual intake.

One reason for the generally modest magnitude of associations between diet quality and overweight and obesity in our study may be because we are examining the two concurrently using cross-sectional data. However, we did find that associations varied by woman’s age with higher GDQS having protective associations among older women, indicating that at the relationship between diet quality and overweight and obesity is likely cumulative. Other studies corroborate these findings. For example, in a longitudinal study in Mexico, non-pregnant, non-lactating women with the largest increase in GDQS over two years had less weight and WC gain, and those with the largest decrease had more weight and WC gain.^48^ Likewise, in the United States, higher GDQS improvement (>5 points) was associated with less weight gain and 23% lower risk of obesity over two and half decades, whereas a higher GDQS decrease (>5 points) was associated with more weight gain and a 32% increase in obesity risk.^37^ At its core, overweight and obesity are fundamentally caused by an imbalance between energy intake and energy expenditure.^49^ The overall small associations between diet quality and overweight and obesity and the large differences in overweight and obesity prevalence by socio-demographic characteristics and location of residence reinforce clinical consensus that overweight and obesity are multifaceted conditions determined by genetics, physiology, psychosocial factors, physical activity and lifestyle, and obesogenic environments.^50^

Our study had several strengths, including the large sample size, rigorous training, and strict operating procedures for anthropometric measurements and dietary assessment. Nevertheless, the study findings should be interpreted with caution for several reasons. First, our analyses were cross-sectional, and none of the results highlighted here are causal. Future research should examine the associations between diet quality and nutritional status longitudinally to establish temporal precedence, causality, and to assess how these associations evolve over time. Second, our sample included WRA who had a biological child between the ages of 10-14 years at enrolment, and findings might not generalise to other WRA or older women. In addition, women excluded from the analysis due to missing data on diet or anthropometry, pregnancy, or very high/low daily energy intake generally came from households where the head was younger and less likely to be female. Lastly, the study sample was not representative of Arusha and Kilimanjaro regions, nor of Tanzania as a whole.

Despite these limitations, our findings can inform the targeting and design of interventions and programmes to prevent and reduce overweight and obesity in Tanzania. The Government of Tanzania already recognises the need to improve the quality of diets and nutritional status of its population.^44,51^ Our research reinforces the need for urgent action to curb the rapidly rising obesity problem, which is likely underestimated given the wide use of BMI as a general obesity metric. Our findings suggest that blanket interventions to improve diet quality alone may have limited success in reducing overweight and obesity in the Tanzanian context. The differences by age group we observed highlight the need for targeted approaches, and suggest that dietary interventions, including those focused on improving diet quality, may be particularly successful among women over 30 years of age, where the overweight and obesity burden was higher and beneficial associations with higher diet quality more pronounced. We also identified several socio-demographic characteristics, including women’s education and occupation, household wealth and food security, and residence in more urbanised villages that could be used to tailor and target obesity prevention and control programs. Rigorous research testing multisectoral obesity prevention and management strategies, including ones to reduce unhealthy diets and reduce physical inactivity,^52,53^ is urgently needed to help curb the obesity pandemic Tanzania faces and reduce the associated rising healthcare burden and costs.

## Data Availability

Data described in the manuscript, code book, and analytic code will be made available by the corresponding author upon request.

## Acknowledgements

We would like to thank respondents for their time and willingness to participate in the study. We acknowledge Neha Kumar for her role in designing the study trial. We are grateful to Gayathri Ramani, Malick Dione, and Rock Zagre for their support in fieldwork preparation, data collection activities, and analytic support for the study. We thank Nishmeet Singh for his support in fieldwork preparation and enumerator training. We acknowledge Nyabasi Makori and Calista N. Njau for their role in processing the diet data. We extend our deepest gratitude to Wiston Mwombeki for his overall support of the study.

## Author contributions

LB, FA, CDA, JK, SYH, and DKO designed the study trial. LB and MTR conceptualised the analyses presented here. LB, FA, and JK oversaw enumerator training. KJ and EM implemented the baseline survey and supervised data collection. LB, FA, and CDA performed data cleaning, processing, and curation. ER conducted the analyses. LB oversaw and validated the analysis. LB drafted the manuscript. All authors interpreted the results and reviewed and approved the final version of the manuscript. LB had the final responsibility for submitting the manuscript for publication.

## Abbreviations

BMI: body mass index
GDQS: Global Diet Quality Score
LMICs: low- and middle-income countries
MD: mean differences
NCDs: non-communicable diseases
PDQS: Prime Dietary Quality Score
RR: risk ratio
WC: waist circumference
WHR: waist-to-hip ratio
WHtR: waist-to-height ratio
WRA: women of reproductive age

