## Supplemental Table for "Diet quality and obesity in women of reproductive age in Northern Tanzania: a cross-sectional study"

**Supplemental Table 1** Scores and number of food items consumed by food groups comprising the Global Diet Quality Score in a sample of 2,415 women of reproductive age

| | Points assigned for consumed amount[1] | | | | | | |
|---|---|---|---|---|---|---|---|
| | **Low** | **Middle** | **High** | **Very high** | **Food group score** | **% who scored 0** | **% who scored the highest** |
| *Healthy* | | | | | | | |
| Citrus fruit | 0 | 1 | 2 | N/A | 0.1±0.3 | 96.9 | 2.4 |
| Deep orange fruit | 0 | 1 | 2 | N/A | 0.1±0.4 | 96.0 | 3.5 |
| Other fruit | 0 | 1 | 2 | N/A | 0.1±0.4 | 94.2 | 3.2 |
| Dark green leafy vegetables | 0 | 2 | 4 | N/A | 2.1±1.9 | 44.5 | 46.9 |
| Cruciferous vegetables | 0 | 0.25 | 0.5 | N/A | 0.2±0.2 | 51.7 | 26.3 |
| Orange vegetables | 0 | 0.25 | 0.5 | N/A | 0.3±0.2 | 12.8 | 39.0 |
| Other vegetables | 0 | 0.25 | 0.5 | N/A | 0.3±0.2 | 18.4 | 32.8 |
| Legumes | 0 | 2 | 4 | N/A | 1.7±1.9 | 52.5 | 37.6 |
| Deep orange tubers | 0 | 0.25 | 0.5 | N/A | 0.0±0.0 | 100.0 | 0.0 |
| Nuts and seeds | 0 | 2 | 4 | N/A | 0.1±0.5 | 97.7 | 1.4 |
| Whole grains | 0 | 1 | 2 | N/A | 1.8±0.5 | 7.8 | 92.2 |
| Liquid oils | 0 | 1 | 2 | N/A | 1.8±0.5 | 5.0 | 85.9 |
| Fish and shellfish | 0 | 1 | 2 | N/A | 0.3±0.6 | 79.9 | 6.2 |
| Poultry and game meat | 0 | 1 | 2 | N/A | 0.0±0.1 | 99.6 | 0.2 |
| Low-fat dairy | 0 | 1 | 2 | N/A | 0.0±0.0 | 99.9 | 0.04 |
| Eggs | 0 | 1 | 2 | N/A | 0.0±0.2 | 97.9 | 1.2 |
| *Unhealthy* | | | | | | | |
| High-fat dairy | 0 | 1 | 2 | 0 | 0.7±0.8 | 55.1 | 23.0 |
| Red meat | 0 | 1 | 0 | N/A | 0.0±0.2 | 96.0 | 4.0 |
| Refined grains and baked foods | 2 | 1 | 0 | N/A | 1.5±0.9 | 26.2 | 73.0 |
| Sweets and ice cream | 2 | 1 | 0 | N/A | 1.0±0.6 | 20.5 | 20.5 |
| Sugar-sweetened beverages | 2 | 1 | 0 | N/A | 1.9±0.4 | 3.6 | 96.4 |
| Juice | 2 | 1 | 0 | N/A | 2.0±0.0 | 0.0 | 100.0 |
| White roots and tubers | 2 | 1 | 0 | N/A | 1.5±0.8 | 20.0 | 66.9 |

| | Points assigned for consumed amount[1] | | | | | | |
|---|---|---|---|---|---|---|---|
| | **Low** | **Middle** | **High** | **Very high** | **Food group score** | **% who scored 0** | **% who scored the highest** |
| Purchased deep fried foods | 2 | 1 | 0 | N/A | 1.5±0.8 | 21.3 | 76.1 |

[1] Intake – Center for Dietary Assessment. (2021). *The Global Diet Quality Score: Data Collection Options and Tabulation Guidelines*. Intake – Center for Dietary Assessment/FHI Solution. https://www.intake.org/sites/default/files/2021-04/GDQS Overview Document - April 2021.pdf

**Supplemental Table 2** Household and individual characteristics of women of reproductive age

| | Included in the analytic sample | Excluded from the analytic sample | p-value for difference |
|---|---|---|---|
| | Mean ± SD or % | Mean ± SD or % | |
| N | 2,415 | 189 | |
| *Household characteristics* | | | |
| Size | 5.8±1.7 | 5.7±1.6 | 0.43 |
| Household head age | 46.8±9.9 | 44.7±9.9 | <0.01 |
| Household head is female | 15.6 | 9.5 | 0.03 |
| Household head has completed primary/higher education | 82.5 | 84.1 | 0.57 |
| *Woman characteristics* | | | |
| Age (in years) | 38.4±6.3 | 36.2±5.9 | <0.01 |
| Has completed primary/higher education | 81.4 | 79.4 | 0.61 |
| Has occupation which requires moderate/heavy physical labour | 63.9 | 60.3 | 0.24 |

**Note:** Women were excluded if they had missing data on diet or anthropometry, they were pregnant at the time of the survey, or had daily energy intake <1$^{st}$ percentile or >99$^{th}$ percentile of the distribution

**Supplemental Table 3** Nutritional status of the 1,933 non-pregnant and non-lactating women of reproductive age (15-49 years of age)

| | **Mean ± SD or N (%)** |
|---|---|
| N | 1,933 |
| Weight (kg) | 67.0±15.6 |
| Height (cm) | 158.9±6.6 |
| Body mass index (BMI, kg/m$^2$) | 26.5±5.9 |
| Hip circumference | 103.0±12.2 |
| Waist circumference | 88.2±13.2 |
| *BMI categories* | |
| Underweight (BMI <18.5 kg/m$^2$) | 4.9 |
| Normal weight (BMI 18.5-24.9 kg/m$^2$) | 40.5 |
| Overweight (BMI 25-29.9 kg/m$^2$) | 29.6 |
| Obesity (BMI 30-34.9 kg/m$^2$) | 16.5 |
| Morbid obesity (BMI≥35 kg/m$^2$) | 8.6 |
| Any overweight/obesity (BMI≥25 kg/m$^2$) | 54.6 |
| General obesity (BMI≥30 kg/m$^2$) | 25.0 |
| *Central obesity* | |
| Waist circumference ≥80 cm | 71.5 |
| Waist circumference ≥88 cm | 48.5 |
| Waist-to-hip ratio ≥0.85 | 53.9 |
| Waist-to-height ratio ≥0.5) | 71.7 |
| Waist-to-hip≥0.85 or BMI≥30 kg/m$^2$ | 60.4 |

**Supplemental Table 4** Nutritional status of the 2,415 women of reproductive age (15-49 years of age) in the analytic sample

| | **Age** | | | | | | **Education** | | | | **Occupation** | | | |
|---|---|---|---|---|---|---|---|---|---|---|---|---|---|---|
| | **20-29 years** | | **30-39 years** | | **40-49 years** | | **No education completed** | | **Primary/higher** | | **Light physical labour required** | | **Moderate/heavy physical labour required** | |
| | **N** | **Mean ± SD or N (%)** | **N** | **Mean ± SD or N (%)** | **N** | **Mean ± SD or N (%)** | **N** | **Mean ± SD or N (%)** | **N** | **Mean ± SD or N (%)** | **N** | **Mean ± SD or N (%)** | **N** | **Mean ± SD or N (%)** |
| Weight (kg) | 183 | 60.2±13.5 | 1136 | 67.6±16.1 | 1096 | 67.2±15.6 | 450 | 59.4±13.3 | 1965 | 68.5±15.8 | 873 | 71.5±17 | 1542 | 64.2±14.5 |
| Body mass index (BMI, kg/m$^2$) | 183 | 23.9±4.9 | 1136 | 26.7±6 | 1096 | 26.6±6 | 450 | 23.6±4.9 | 1965 | 27.1±6 | 873 | 28.4±6.4 | 1542 | 25.4±5.4 |
| Hip circumference (cm) | 183 | 98.0±11.8 | 1136 | 103.4±12.1 | 1096 | 103.1±12.3 | 450 | 97±11.1 | 1965 | 104.2±12.1 | 873 | 106.5±13.1 | 1542 | 100.8±11.2 |
| Waist circumference (cm) | 183 | 82.1±12.7 | 1136 | 87.9±13 | 1096 | 89.3±13.4 | 450 | 82.5±12.3 | 1965 | 89.4±13.2 | 873 | 91.6±13.8 | 1542 | 86.2±12.6 |
| *BMI categories* | | | | | | | | | | | | | | |
| Underweight (BMI <18.5 kg/m$^2$) | 183 | 17 (9.3) | 1136 | 49 (4.3) | 1096 | 51 (4.7) | 450 | 44 (9.8) | 1965 | 73 (3.7) | 873 | 29 (3.3) | 1542 | 88 (5.7) |
| Normal weight (BMI 18.5-24.9 kg/m$^2$) | 183 | 102 (55.7) | 1136 | 467 (41.1) | 1096 | 435 (39.7) | 450 | 265 (58.9) | 1965 | 739 (37.6) | 873 | 265 (30.4) | 1542 | 739 (47.9) |
| Overweight (BMI 25-29.9 kg/m$^2$) | 183 | 42 (23) | 1136 | 318 (28) | 1096 | 329 (30) | 450 | 90 (20) | 1965 | 599 (30.5) | 873 | 261 (29.9) | 1542 | 428 (27.8) |
| Obesity (BMI 30-34.9 kg/m$^2$) | 183 | 15 (8.2) | 1136 | 197 (17.3) | 1096 | 190 (17.3) | 450 | 35 (7.8) | 1965 | 367 (18.7) | 873 | 197 (22.6) | 1542 | 205 (13.3) |
| Morbid obesity (BMI≥35 kg/m$^2$) | 183 | 7 (3.8) | 1136 | 105 (9.2) | 1096 | 91 (8.3) | 450 | 16 (3.6) | 1965 | 187 (9.5) | 873 | 121 (13.9) | 1542 | 82 (5.3) |
| Any overweight/obesity (BMI≥25 kg/m$^2$) | 183 | 64 (35) | 1136 | 620 (54.6) | 1096 | 610 (55.7) | 450 | 141 (31.3) | 1965 | 1153 (58.7) | 873 | 579 (66.3) | 1542 | 715 (46.4) |
| General obesity (BMI≥30 kg/m$^2$) | 183 | 22 (12) | 1136 | 302 (26.6) | 1096 | 281 (25.6) | 450 | 51 (11.3) | 1965 | 554 (28.2) | 873 | 318 (36.4) | 1542 | 287 (18.6) |
| *Central obesity* | | | | | | | | | | | | | | |
| Waist circumference ≥80 cm | 183 | 95 (51.9) | 1136 | 811 (71.4) | 1096 | 810 (73.9) | 450 | 232 (51.6) | 1965 | 1484 (75.5) | 873 | 685 (78.5) | 1542 | 1031 (66.9) |
| Waist circumference ≥88 cm | 183 | 50 (27.3) | 1136 | 548 (48.2) | 1096 | 566 (51.6) | 450 | 142 (31.6) | 1965 | 1022 (52) | 873 | 531 (60.8) | 1542 | 633 (41.1) |
| Waist-to-hip ratio ≥0.85 | 183 | 76 (41.5) | 1136 | 566 (49.8) | 1096 | 652 (59.5) | 450 | 232 (51.6) | 1965 | 1062 (54) | 873 | 484 (55.4) | 1542 | 810 (52.5) |

| | | | | | | | | | | | | |
|---|---|---|---|---|---|---|---|---|---|---|---|---|
| Waist-to-height ratio ≥0.5 | 183 | 104 (56.8) | 1136 | 816 (71.8) | 1096 | 808 (73.7) | 450 | 247 (54.9) | 1965 | 1481 (75.4) | 873 | 701 (80.3) | 1542 | 1027 (66.6) |
| Waist-to-hip ratio ≥0.85 or BMI≥30 $kg/m^2$ | 183 | 82 (44.8) | 1136 | 660 (58.1) | 1096 | 717 (65.4) | 450 | 239 (53.1) | 1965 | 1220 (62.1) | 873 | 582 (66.7) | 1542 | 877 (56.9) |

| | Wealth quintile | | | | | | | | | |
|---|---|---|---|---|---|---|---|---|---|---|
| | **1st quintile** | | **2nd quintile** | | **3rd quintile** | | **4th quintile** | | **5th quintile** | |
| | **N** | **Mean ± SD or N (%)** | **N** | **Mean ± SD or N (%)** | **N** | **Mean ± SD or N (%)** | **N** | **Mean ± SD or N (%)** | **N** | **Mean ± SD or N (%)** |
| Weight (kg) | 475 | 57.9±12.3 | 486 | 62.8±13.3 | 488 | 67.1±15.5 | 485 | 71±16 | 481 | 75.4±15.4 |
| Body mass index (BMI, $kg/m^2$) | 475 | 23±4.6 | 486 | 24.9±5.1 | 488 | 26.6±5.9 | 485 | 28±6 | 481 | 29.8±5.8 |
| Hip circumference (cm) | 475 | 95.9±10.1 | 486 | 99.5±10.6 | 488 | 103.1±11.8 | 485 | 105.9±11.5 | 481 | 109.9±12.1 |
| Waist circumference (cm) | 475 | 80.8±11.3 | 486 | 85.5±11.8 | 488 | 88.4±13 | 485 | 91.3±13 | 481 | 94.5±13 |
| *BMI categories* | | | | | | | | | | |
| Underweight (BMI <18.5 $kg/m^2$) | 475 | 55 (11.6) | 486 | 31 (6.4) | 488 | 17 (3.5) | 485 | 11 (2.3) | 481 | 3 (0.6) |
| Normal weight (BMI 18.5-24.9 $kg/m^2$) | 475 | 304 (64) | 486 | 253 (52.1) | 488 | 196 (40.2) | 485 | 150 (30.9) | 481 | 101 (21) |
| Overweight (BMI 25-29.9 $kg/m^2$) | 475 | 71 (14.9) | 486 | 130 (26.7) | 488 | 155 (31.8) | 485 | 169 (34.8) | 481 | 164 (34.1) |
| Obesity (BMI 30-34.9 $kg/m^2$) | 475 | 38 (8) | 486 | 51 (10.5) | 488 | 80 (16.4) | 485 | 100 (20.6) | 481 | 133 (27.7) |
| Morbid obesity (BMI≥35 $kg/m^2$) | 475 | 7 (1.5) | 486 | 21 (4.3) | 488 | 40 (8.2) | 485 | 55 (11.3) | 481 | 80 (16.6) |
| Any overweight/obesity (BMI≥25 $kg/m^2$) | 475 | 116 (24.4) | 486 | 202 (41.6) | 488 | 275 (56.4) | 485 | 324 (66.8) | 481 | 377 (78.4) |
| General obesity (BMI≥30 $kg/m^2$) | 475 | 45 (9.5) | 486 | 72 (14.8) | 488 | 120 (24.6) | 485 | 155 (32) | 481 | 213 (44.3) |
| *Central obesity* | | | | | | | | | | |
| Waist circumference ≥80 cm | 475 | 224 (47.2) | 486 | 312 (64.2) | 488 | 356 (73) | 485 | 397 (81.9) | 481 | 427 (88.8) |
| Waist circumference ≥88 cm | 475 | 110 (23.2) | 486 | 188 (38.7) | 488 | 236 (48.4) | 485 | 288 (59.4) | 481 | 342 (71.1) |
| Waist-to-hip ratio ≥0.85 | 475 | 219 (46.1) | 486 | 264 (54.3) | 488 | 264 (54.1) | 485 | 281 (57.9) | 481 | 266 (55.3) |
| Waist-to-height ratio ≥0.5 | 475 | 245 (51.6) | 486 | 317 (65.2) | 488 | 347 (71.1) | 485 | 396 (81.6) | 481 | 423 (87.9) |
| Waist-to-hip ratio ≥0.85 or BMI≥30 $kg/m^2$ | 475 | 228 (48) | 486 | 279 (57.4) | 488 | 295 (60.5) | 485 | 315 (64.9) | 481 | 342 (71.1) |

| | Household food insecurity | | | | | | | | Urbanicity tertile | | | | | |
|---|---|---|---|---|---|---|---|---|---|---|---|---|---|---|
| | None | | Mild | | Moderate | | Severe | | Lower (rural) | | Middle (peri-urban) | | Upper (urban) | |
| | N | Mean ± SD or N (%) | N | Mean ± SD or N (%) | N | Mean ± SD or N (%) | N | Mean ± SD or N (%) | N | Mean ± SD or N (%) | N | Mean ± SD or N (%) | N | Mean ± SD or N (%) |
| Weight (kg) | 871 | 70.9±16.6 | 357 | 66.8±14.7 | 409 | 66.6±15.3 | 778 | 62.5±14.5 | 814 | 60.5±13.6 | 820 | 68±15.1 | 781 | 72.2±16.4 |
| Body mass index (BMI, kg/m$^2$) | 871 | 27.9±6.2 | 357 | 26.7±5.9 | 409 | 26.4±5.8 | 778 | 24.8±5.4 | 814 | 24±5 | 820 | 26.8±5.7 | 781 | 28.8±6.2 |
| Hip circumference (cm) | 871 | 105.7±12.5 | 357 | 103.3±12.2 | 409 | 102.5±11.9 | 778 | 99.7±11.4 | 814 | 97.7±10.5 | 820 | 103.9±11.5 | 781 | 107.2±12.8 |
| Waist circumference (cm) | 871 | 90.8±13.5 | 357 | 88±13 | 409 | 88.3±13.2 | 778 | 85±12.6 | 814 | 82.9±11.4 | 820 | 89.1±12.8 | 781 | 92.5±13.7 |
| *BMI categories* | | | | | | | | | | | | | | |
| Underweight (BMI <18.5 kg/m$^2$) | 871 | 29 (3.3) | 357 | 12 (3.4) | 409 | 14 (3.4) | 778 | 62 (8) | 814 | 64 (7.9) | 820 | 33 (4) | 781 | 20 (2.6) |
| Normal weight (BMI 18.5-24.9 kg/m$^2$) | 871 | 284 (32.6) | 357 | 145 (40.6) | 409 | 184 (45) | 778 | 391 (50.3) | 814 | 477 (58.6) | 820 | 317 (38.7) | 781 | 210 (26.9) |
| Overweight (BMI 25-29.9 kg/m$^2$) | 871 | 267 (30.7) | 357 | 111 (31.1) | 409 | 111 (27.1) | 778 | 200 (25.7) | 814 | 185 (22.7) | 820 | 248 (30.2) | 781 | 256 (32.8) |
| Obesity (BMI 30-34.9 kg/m$^2$) | 871 | 189 (21.7) | 357 | 57 (16) | 409 | 65 (15.9) | 778 | 91 (11.7) | 814 | 67 (8.2) | 820 | 154 (18.8) | 781 | 181 (23.2) |
| Morbid obesity (BMI≥35 kg/m$^2$) | 871 | 102 (11.7) | 357 | 32 (9) | 409 | 35 (8.6) | 778 | 34 (4.4) | 814 | 21 (2.6) | 820 | 68 (8.3) | 781 | 114 (14.6) |
| Any overweight/obesity (BMI≥25 kg/m$^2$) | 871 | 558 (64.1) | 357 | 200 (56) | 409 | 211 (51.6) | 778 | 325 (41.8) | 814 | 273 (33.5) | 820 | 470 (57.3) | 781 | 551 (70.6) |
| General obesity (BMI≥30 kg/m$^2$) | 871 | 291 (33.4) | 357 | 89 (24.9) | 409 | 100 (24.4) | 778 | 125 (16.1) | 814 | 88 (10.8) | 820 | 222 (27.1) | 781 | 295 (37.8) |
| *Central obesity* | | | | | | | | | | | | | | |
| Waist circumference ≥80 cm | 871 | 689 (79.1) | 357 | 251 (70.3) | 409 | 291 (71.1) | 778 | 485 (62.3) | 814 | 451 (55.4) | 820 | 631 (77) | 781 | 634 (81.2) |
| Waist circumference ≥88 cm | 871 | 505 (58) | 357 | 164 (45.9) | 409 | 197 (48.2) | 778 | 298 (38.3) | 814 | 251 (30.8) | 820 | 418 (51) | 781 | 495 (63.4) |
| Waist-to-hip ratio ≥0.85 | 871 | 480 (55.1) | 357 | 183 (51.3) | 409 | 231 (56.5) | 778 | 400 (51.4) | 814 | 388 (47.7) | 820 | 462 (56.3) | 781 | 444 (56.9) |

| | | | | | | | | | | | | | | |
|---|---|---|---|---|---|---|---|---|---|---|---|---|---|---|
| Waist-to-height ratio ≥0.5 | 871 | 687 (78.9) | 357 | 251 (70.3) | 409 | 297 (72.6) | 778 | 493 (63.4) | 814 | 479 (58.8) | 820 | 608 (74.1) | 781 | 641 (82.1) |
| Waist-to-hip ratio ≥0.85 or BMI≥30 kg/m$^2$ | 871 | 568 (65.2) | 357 | 203 (56.9) | 409 | 259 (63.3) | 778 | 429 (55.1) | 814 | 415 (51) | 820 | 512 (62.4) | 781 | 532 (68.1) |

**Supplemental Table 5** Bivariate and multivariable adjusted associations between diet quality and nutritional status in 2,415 women of reproductive age (binary outcomes)

| | | GDQS | GDQS+ | GDQS- |
|---|---|---|---|---|
| Underweight (BMI <18.5 kg/m$^2$) | Unadjusted | 1.02 (0.98, 1.06) | 0.97 (0.92, 1.02) | 1.2 (1.09, 1.32) |
| | Adjusted | 0.98 (0.94, 1.02) | 0.97 (0.92, 1.02) | 1.01 (0.93, 1.11) |
| Normal weight (BMI 18.5-24.9 kg/m$^2$) | Unadjusted | 1.03 (1.02, 1.05) | 1.02 (1, 1.04) | 1.09 (1.06, 1.13) |
| | Adjusted | 1.01 (1, 1.02) | 1.01 (1, 1.03) | 1.01 (0.98, 1.04) |
| Any overweight/obesity (BMI≥25 kg/m$^2$) | Unadjusted | 0.97 (0.96, 0.98) | 0.99 (0.97, 1) | 0.92 (0.89, 0.95) |
| | Adjusted | 1 (0.99, 1.01) | 1 (0.98, 1.01) | 0.99 (0.98, 1.01) |
| General obesity (BMI≥30 kg/m$^2$) | Unadjusted | 0.95 (0.93, 0.97) | 0.97 (0.95, 0.99) | 0.9 (0.86, 0.95) |
| | Adjusted | 0.99 (0.97, 1.01) | 0.98 (0.96, 1) | 1.01 (0.98, 1.04) |
| Morbid obesity (BMI≥35 kg/m$^2$) | Unadjusted | 0.92 (0.89, 0.95) | 0.93 (0.89, 0.97) | 0.9 (0.83, 0.97) |
| | Adjusted | 0.97 (0.94, 1) | 0.95 (0.91, 0.99) | 1.03 (0.97, 1.1) |
| *Central obesity* | | | | |
| Waist circumference ≥80 cm | Unadjusted | 0.99 (0.98, 1) | 1 (0.99, 1.01) | 0.96 (0.94, 0.98) |
| | Adjusted | 1 (1, 1.01) | 1.01 (1, 1.02) | 1 (0.98, 1.01) |
| Waist circumference ≥88 cm | Unadjusted | 0.97 (0.96, 0.99) | 0.99 (0.97, 1) | 0.93 (0.9, 0.96) |
| | Adjusted | 1 (0.98, 1.01) | 1 (0.98, 1.01) | 1 (0.98, 1.01) |
| Waist-to-hip ratio ≥0.85 | Unadjusted | 1 (0.99, 1.01) | 1 (0.99, 1.01) | 0.99 (0.97, 1.01) |
| | Adjusted | 1 (0.99, 1.02) | 1 (0.99, 1.02) | 1.01 (0.99, 1.02) |
| Waist-to-height ratio ≥0.5 | Unadjusted | 0.99 (0.98, 1) | 1 (0.99, 1.01) | 0.95 (0.94, 0.97) |
| | Adjusted | 1 (0.99, 1.01) | 1 (0.99, 1.01) | 0.99 (0.98, 1) |
| Waist-to-hip ratio ≥0.85 or BMI≥30 kg/m$^2$ | Unadjusted | 0.99 (0.98, 1) | 0.99 (0.98, 1.01) | 0.98 (0.95, 1) |
| | Adjusted | 1 (0.99, 1.01) | 1 (0.98, 1.01) | 1.01 (0.99, 1.02) |

**Note:** All values are risk ratios (RR) and 95% CI from generalised linear models. Adjusted estimates control for woman's age, education, occupation, household wealth, region, and urbanicity. Standard errors were clustered at the village level. Abbreviations used: BMI, body mass index; GDQS, global diet quality score

**Supplemental Table 6** Bivariate and multivariable adjusted associations between diet quality and nutritional status in 1,933 non-pregnant and non-lactating women of reproductive age (binary outcomes)

| | | GDQS | GDQS+ | GDQS- |
|---|---|---|---|---|
| Underweight (BMI <18.5 kg/m$^2$) | Unadjusted | 1.02 (0.98, 1.07) | 0.98 (0.91, 1.04) | 1.2 (1.07, 1.34) |
| | Adjusted | 0.98 (0.94, 1.03) | 0.97 (0.92, 1.03) | 1.03 (0.92, 1.15) |
| Normal weight (BMI 18.5-24.9 kg/m$^2$) | Unadjusted | 1.04 (1.02, 1.05) | 1.02 (1, 1.04) | 1.1 (1.05, 1.14) |
| | Adjusted | 1.01 (1, 1.03) | 1.01 (0.99, 1.03) | 1.02 (0.98, 1.05) |
| Any overweight/obesity (BMI≥25 kg/m$^2$) | Unadjusted | 0.97 (0.96, 0.99) | 0.99 (0.97, 1.01) | 0.92 (0.89, 0.96) |
| | Adjusted | 0.99 (0.98, 1) | 1 (0.99, 1.01) | 0.99 (0.97, 1) |
| General obesity (BMI≥30 kg/m$^2$) | Unadjusted | 0.96 (0.94, 0.98) | 0.97 (0.95, 1) | 0.91 (0.86, 0.97) |
| | Adjusted | 0.99 (0.97, 1.01) | 0.99 (0.96, 1.01) | 1.01 (0.98, 1.04) |
| Morbid obesity (BMI≥35 kg/m$^2$) | Unadjusted | 0.93 (0.89, 0.96) | 0.93 (0.88, 0.97) | 0.91 (0.84, 1) |
| | Adjusted | 0.97 (0.93, 1.0079) | 0.95 (0.9, 1) | 1.03 (0.96, 1.11) |
| *Central obesity* | | | | |
| Waist circumference ≥80 cm | Unadjusted | 0.99 (0.98, 1) | 1 (0.99, 1.02) | 0.96 (0.93, 0.98) |
| | Adjusted | 1 (1, 1.01) | 1.01 (1, 1.02) | 0.99 (0.98, 1.01) |
| Waist circumference ≥88 cm | Unadjusted | 0.97 (0.96, 0.99) | 0.99 (0.97, 1.01) | 0.93 (0.9, 0.96) |
| | Adjusted | 1 (0.98, 1.01) | 1 (0.98, 1.01) | 0.99 (0.97, 1.01) |
| Waist-to-hip ratio ≥0.85 | Unadjusted | 1 (0.98, 1.01) | 1 (0.98, 1.01) | 0.99 (0.97, 1.02) |
| | Adjusted | 1 (0.99, 1.02) | 1 (0.99, 1.02) | 1.01 (0.99, 1.03) |
| Waist-to-height ratio ≥0.5 | Unadjusted | 0.99 (0.98, 1) | 1 (0.99, 1.01) | 0.95 (0.94, 0.97) |
| | Adjusted | 1 (0.99, 1.01) | 1 (0.99, 1.01) | 0.99 (0.97, 1) |
| Waist-to-hip ratio ≥0.85 or BMI≥30 kg/m$^2$ | Unadjusted | 0.99 (0.98, 1) | 0.99 (0.98, 1.01) | 0.98 (0.95, 1) |
| | Adjusted | 1 (0.99, 1.01) | 1 (0.98, 1.01) | 1.01 (0.99, 1.02) |

**Note:** All values are risk ratios (RR) and 95% CI from generalised linear models. Adjusted estimates control for woman's age, education, occupation, household wealth, region, and urbanicity. Standard errors were clustered at the village level. Abbreviations used: BMI, body mass index; GDQS, global diet quality score

**Supplemental Table 7** Bivariate and multivariable adjusted associations between diet quality and nutritional status in 2,415 women of reproductive age (continuous outcomes)

| | | **GDQS** | **GDQS+** | **GDQS-** |
|---|---|---|---|---|
| BMI (kg/m$^2$) | Unadjusted | -0.22 (-0.29, -0.15) | -0.12 (-0.21, -0.03) | -0.54 (-0.74, -0.35) |
| | Adjusted | -0.06 (-0.12, 0.01) | -0.07 (-0.14, 0.01) | -0.01 (-0.13, 0.11) |
| Waist circumference (cm) | Unadjusted | -0.4 (-0.56, -0.23) | -0.18 (-0.39, 0.02) | -1.08 (-1.5, -0.65) |
| | Adjusted | -0.08 (-0.22, 0.07) | -0.09 (-0.25, 0.08) | -0.04 (-0.31, 0.23) |
| Hip circumference (cm) | Unadjusted | -0.42 (-0.57, -0.27) | -0.19 (-0.35, -0.03) | -1.14 (-1.51, -0.77) |
| | Adjusted | -0.1 (-0.23, 0.02) | -0.11 (-0.23, 0.02) | -0.09 (-0.36, 0.17) |

**Note:** All values are mean differences (MD) and 95% CI from generalised linear models. Adjusted estimates control for woman's age, education, occupation, household wealth, region, and urbanicity. Standard errors were clustered at the village level. Abbreviations used: BMI, body mass index; GDQS, global diet quality score.

**Supplemental Table 8** Bivariate and multivariable adjusted associations between diet quality and nutritional status in 1,933 non-pregnant and non-lactating women of reproductive age (continuous outcomes)

| | | GDQS | GDQS+ | GDQS- |
|---|---|---|---|---|
| BMI (kg/m$^2$) | Unadjusted | -0.21 (-0.29, -0.14) | -0.11 (-0.2, -0.02) | -0.54 (-0.75, -0.32) |
| | Adjusted | -0.06 (-0.12, 0.003) | -0.06 (-0.14, 0.02) | -0.05 (-0.18, 0.07) |
| Waist circumference (cm) | Unadjusted | -0.37 (-0.54, -0.19) | -0.16 (-0.38, 0.06) | -1.02 (-1.5, -0.55) |
| | Adjusted | -0.07 (-0.22, 0.08) | -0.07 (-0.26, 0.12) | -0.06 (-0.36, 0.23) |
| Hip circumference (cm) | Unadjusted | -0.37 (-0.53, -0.2) | -0.13 (-0.31, 0.04) | -1.12 (-1.51, -0.73) |
| | Adjusted | -0.08 (-0.21, 0.04) | -0.06 (-0.2, 0.08) | -0.18 (-0.44, 0.09) |

**Note:** All values are mean differences (MD) and 95% CI from generalised linear models. Adjusted estimates control for woman's age, education, occupation, household wealth, region, and urbanicity. Standard errors were clustered at the village level. Abbreviations used: BMI, body mass index; GDQS, global diet quality score.

**Supplemental Table 9** Heterogeneity of the associations between total GDQS and nutritional status by woman's age group in 1,933 non-pregnant and non-lactating women of reproductive age

| | **20-29 years old** | **30-39 years old** | **40-49 years old** |
|---|---|---|---|
| General obesity (BMI≥30 kg/m$^2$) | 1.05 (.099, 1.12) | 0.92 (0.86, 0.98) | 0.95 (0.89, 1.02) |
| Morbid obesity (BMI≥35 kg/m$^2$) | 1.07 (0.93, 1.24) | 0.91 (0.78, 1.06) | 0.86 (0.77, 1.02) |
| Waist circumference ≥88 cm | 1.05 (0.99, 1.11) | 0.94 (0.89, 0.99) | 0.95 (0.90, 1.01) |
| BMI (kg/m$^2$) | 0.19 (0.06, 0.32) | -0.27 (-0.43, -0.11) | -0.25 (-0.42, -0.09) |
| Waist circumference (cm) | 0.48 (0.18, 0.79) | -0.66 (-1.03, -0.29) | -0.54 (-0.91, -0.17) |
| Hip circumference (cm) | 0.41 (-0.03, 0.84) | -0.58 (-1.06, -0.09) | -0.53 (-1.00, -0.05) |

**Note:** All values are mean differences (MD) and 95% CI from generalised linear models. Adjusted estimates control for woman's age, education, occupation, household wealth, region, and urbanicity. Standard errors were clustered at the village level. Abbreviations used: BMI, body mass index; GDQS, global diet quality score.
